# The adoption and implementation of local government planning regulations to manage hot food takeaways near schools in England: A qualitative process evaluation

**DOI:** 10.1101/2024.07.18.24310617

**Authors:** S Hassan, C Thompson, J Adams, M Chang, D Derbyshire, M Keeble, B Liu, OT Mytton, J Rahilly, B Savory, R Smith, M White, T Burgoine, S Cummins

**Author notes:** UCL Great Ormond Street Institute of Child Health, 30 Guilford Street London WC1N 1EH, UK.

## Abstract

**Introduction:** Access to hot food takeaways, particularly near schools, is of growing concern for policymakers seeking to reduce childhood obesity globally. In England, United Kingdom (UK), local government jurisdictions are implementing planning policies to reduce access by restricting or denying planning permission for new takeaway outlets near schools. We used a qualitative approach to explore local government officers’ perspectives on the barriers to and facilitators of the adoption, implementation, and perceived effectiveness of these policies.

**Methods:** In 2021-2022, we conducted semi-structured interviews with 29 local planning (‘planners’) and public health government officers from 15 different local authorities across England who adopted a policy to restrict new takeaways. Data were analysed thematically.

**Results:** Participants explained that they mostly thought the policies facilitated the refusal of applications for new takeaways near schools. However, participants speculated that businesses identified alternative opportunities to operate including functioning as ‘restaurants’ or within other locations. Effective working relationships between planners and public health officers were important for adoption and implementation, although planning and public health agendas did not always align and there were tensions between economic development and health improvement goals. The policy was adapted to suit local needs and priorities; in some cases, the policy was not used in areas where economic growth was prioritised. Clarity in policy wording and establishing a formal process for implementing policies including a designated individual responsible for checking and reviewing takeaway applications helped ensure consistency and confidence in policy implementation.

**Conclusion:** Although sometimes challenging, the policies were commonly described as feasible to implement. However, they may not completely prevent new takeaways opening, particularly where takeaways are relied upon to enhance local economies or where takeaway businesses find alternative ways to operate. Nevertheless, the policies can serve to shift the balance of power that currently favours commercial interests over public health priorities.

**Highlights:**

- Planning and public health officers struggled to align economic and health agendas
- Policy champions helped align agendas and push takeaway management policies forward
- Policies were adapted to avoid use where they negatively impacted economic growth
- Established processes and clearly worded policies facilitated policy implementation
- The polices made it easier to deny planning permission for new takeaways

## Introduction

Hot food takeaways are outlets that typically sell food to be consumed away from the premises. Other terms used in the literature include ‘fast food’ and ‘takeout’, however we use the term ‘takeaways’ throughout. The global takeaways industry reached an estimated revenue of $978.4 billion in 2023, with an annual growth rate of 2.1% over five years prior to 2023 and is projected to grow further (IBISWorld, 2023). The demand for takeaways is therefore increasing globally.

Takeaways typically sell foods that are high in total energy, fat, salt, sugar and calories which contribute toward weight gain, diet-related health problems in children and adults globally (Davies et al., 2016; Donin et al., 2018; Filgueiras et al., 2023; Huang et al., 2021; Jaworowska et al., 2014; Jia et al., 2023; Jiang et al., 2023; Ntarladima et al., 2022). There is evidence that that takeaways are more prevalent in deprived areas and near schools in Hong Kong, Australia and England, UK (Cheung et al., 2021; Maguire et al., 2015; Smith et al., 2013; Trapp et al., 2022; Turbutt et al., 2019). There is additionally global evidence for an association between physical exposure to takeaways, takeaway food consumption, and obesity (Burgoine et al., 2016; Burgoine et al., 2014; Burgoine et al., 2018; Filgueiras et al., 2023; Jia et al., 2023; Jiang et al., 2023; Ohri-Vachaspati et al., 2023).

Takeaways are a public health concern among policy makers targeting the prevention of obesity in children. The World Health Organisation advocates strategies to prevent the sale of products high in fats and sugars near schools (World Health Organisation (WHO), 2022). One method is making using of the urban planning system (i.e., whereby developments, built environments and land use are actively managed to meet community and political needs) to reduce the availability of takeaways. An early example of this was in Los Angeles, USA, where in 2008 new ‘fast food’ outlets were restricted within targeted zones (Los Angeles City Planning, 2007; Sturm & Hattori, 2015). Similarly, drive-through fast food takeaways were restricted across Western and Eastern parts of Canada (Nykiforuk et al., 2018). There has also been a focus on food environments around schools; for example, Ireland introduced a ‘no fry zone’ specifically restricting takeaways within 400 metres from schools or playgrounds (Harrington et al., 2020; Moyles, 2018).

In 2019, 41 LAs (from 325) LAs in England adopted policies and/or planning guidance whereby applications for new takeaways could be denied planning permission if they fell within a specific distance of a school (Keeble et al., 2024 (in preparation)) . In the England, opening a new takeaway requires planning permission from the relevant local government jurisdiction (i.e. ‘local authority’ (LA)) (Ministry of Housing Communities and Local Government, 2019). According to current planning guidelines, new takeaways are those opening in new retail units, as well as those opening in premises in existing buildings where the previous retail use was not a takeaway (i.e. change of use). Before September 2020, takeaways were categorised separately from other types of non-residential buildings as category “A5” (Town and Country Planning England, 2005). After September 2020, the A5 classification was replaced with the category “*sui generis*” (literally “of its own kind”). The separation of this category of non-residential buildings may prove advantageous in attempting to curate a healthy food environment as takeaways can be specifically targeted (Public Health England, 2021).

Polices restricting new takeaways opening within certain distances of schools in England are commonly known as “exclusion zones” (Keeble et al., 2021). Although these policies cannot be applied retrospectively to close existing takeaways and does not apply to renewals or opening another takeaway on the site of an old one, it permits refusal of, or restricted planning permission for, *new* takeaways near schools. We use the term “takeaway management zones” rather than exclusion zones to reflect the variation in approaches adopted by LAs. Most management zones use a 400-metre Euclidean distance from the boundary of a school (i.e. where planning permission is refused if falling within this metric), which is deemed to be roughly equal to a five minute walk (see Figure 1 for variations) (Rahilly et al., 2024). Some policies focus on secondary schools (children aged 11-16 years old) only, whilst others include both primary (children aged 5-11 years old) and secondary schools. Others may also exclude town centres (i.e., areas defined by LAs and includes location of retail, commercial, leisure and cultural uses) where these overlap management zones. In some instances, new takeaways are required to restrict their hours of operation.

**Figure 1.**
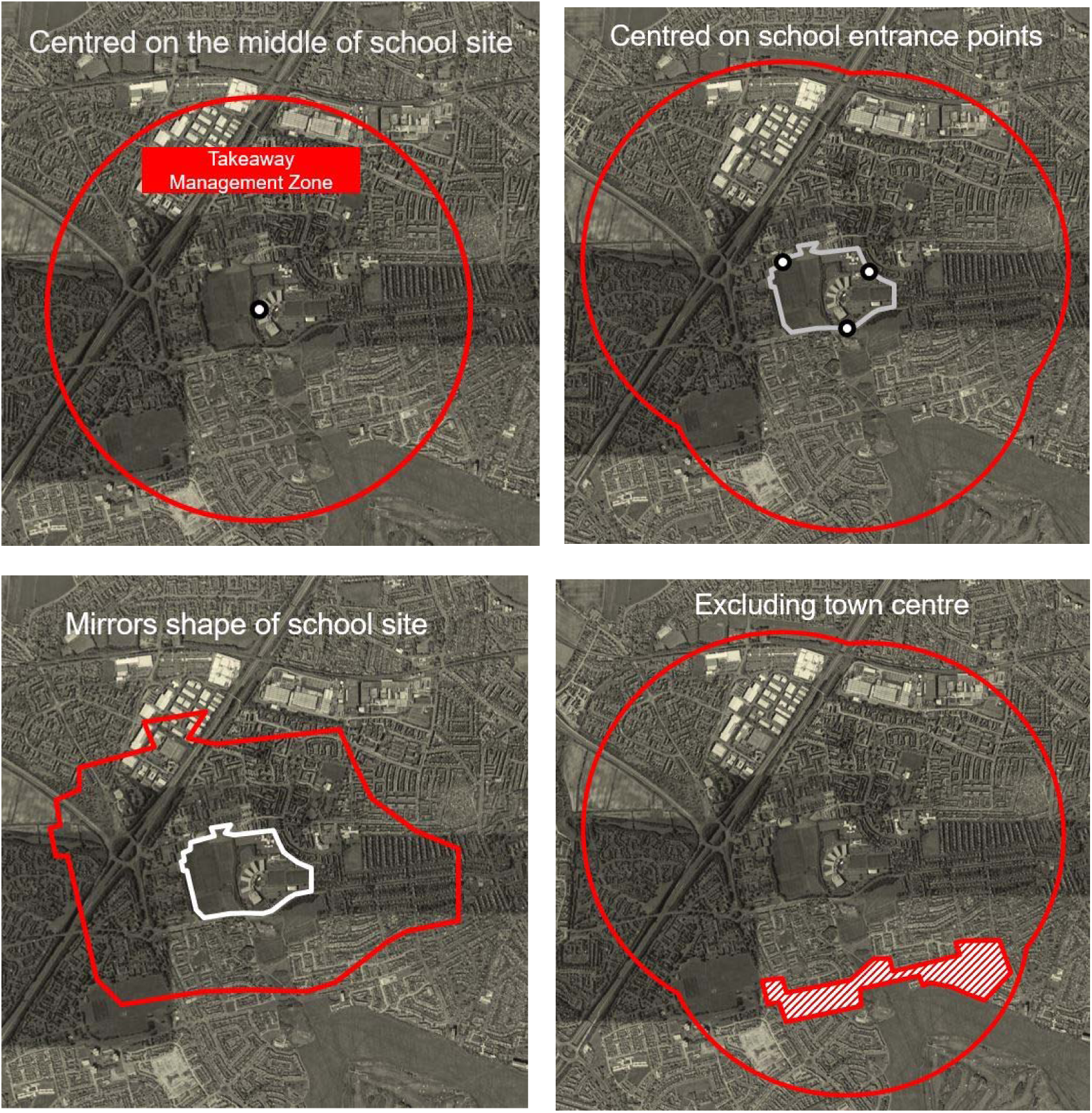
Aerial views demonstrating variation in the types of takeaway management zones adopted by LAs in England, UK.

The processes involved in the adoption (i.e., creating policies and guidance, and obtaining relevant approvals) and implementation (i.e. putting into effect policies to deny or restrict planning applications) of takeaway management zones are displayed in Figure 2. For LA planners to review takeaway planning applications against takeaway management zones, they must first be adopted as policies within a local plan (i.e., statutory document outlining the future development of an area through the adoption of planning policies). Takeaway management zones may not be referenced within a local plan, although they can be, but there must be a broader health policy within the local plan. Supplementary Planning Documents serve as further guidance to explain policies in the local plan and may include further guidance on takeaway management zones but cannot be used to make new policies.

**Figure 2.**
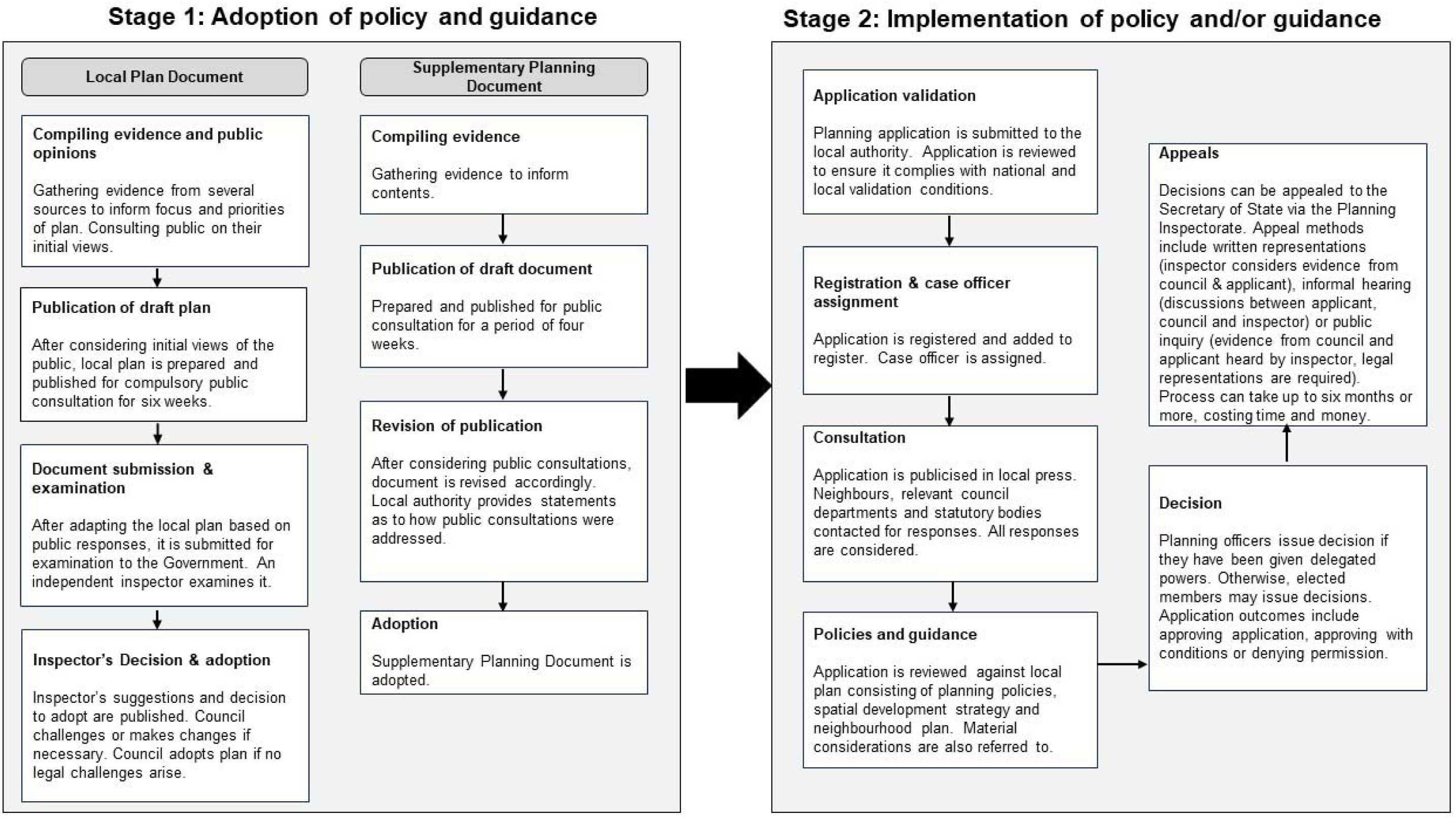
A summary of the process of adopting and implementing planning policies and guidance within LAs in England, UK* (The Town and Country Planning (Local Planning) (England) Regulations., 2012; Town Planning Info a; Town Planning Info b) *See box 1 for definition of key terms

### Box 1.

Definition of key terms described in *Figure 2*.

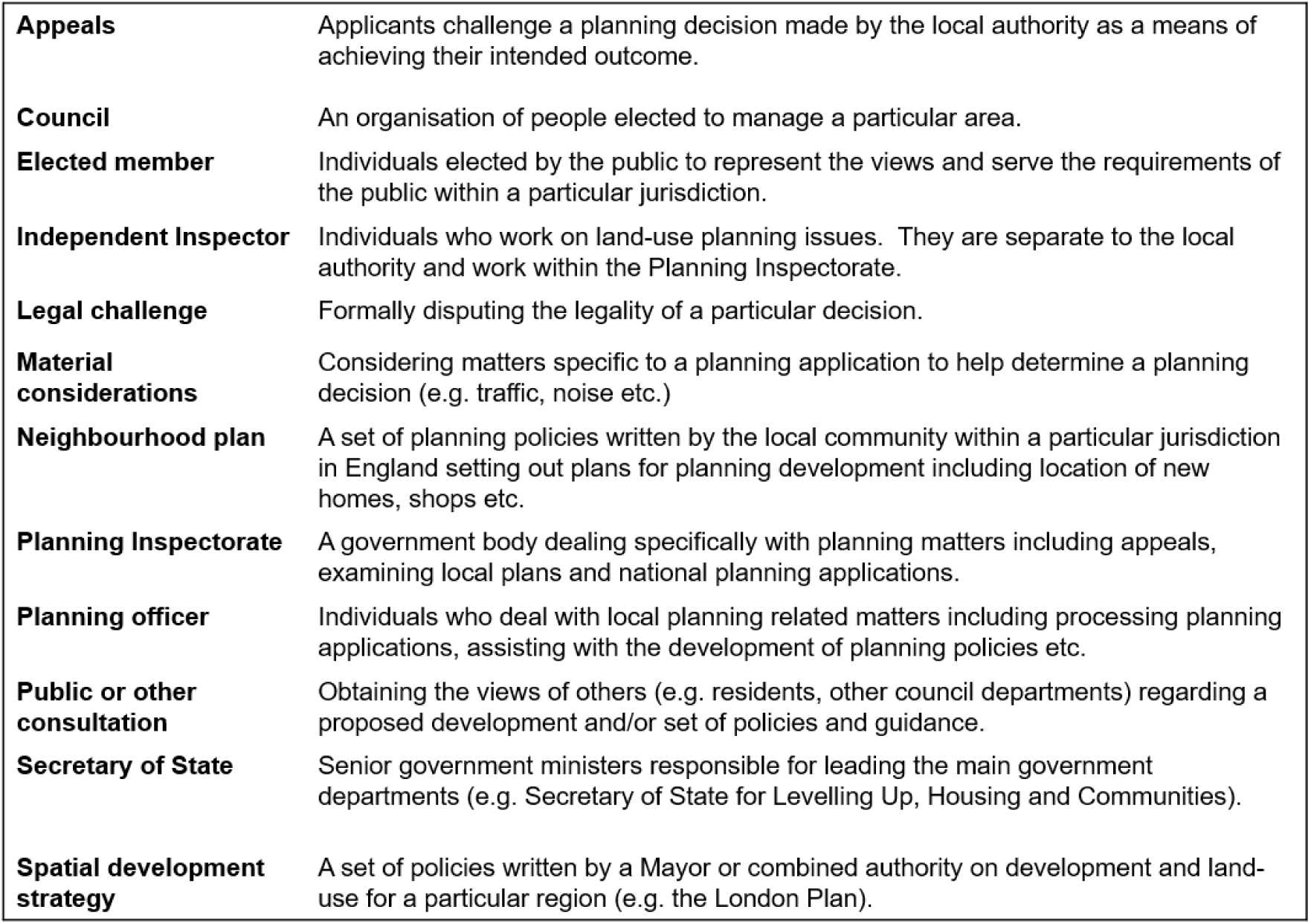

The different takeaway management approaches adopted by LAs and the different local contexts in which they are implemented raises questions around how best to go about operationalising these policies and the practical issues they present. Yet there is limited research investigating the adoption and implementation of takeaway management zones near schools or other takeaway restriction policies both in England and internationally. It is yet to be determined if particular approaches and variations are perceived to be more successful than others (Keeble et al., 2021). Improving understanding of context may aid understanding of why food environment policies succeed or fail, and how adaptations play-out across different settings will help to inform practice for countries seeking to adopt and implement such policies (Mah et al., 2019). Practitioners seeking to implement food environment policies face a common set of barriers, including lack of stakeholder engagement or prioritisation of policy, resistance to change, and concern over commercial revenue (Nguyen et al., 2021). Improving understanding of how these barriers are experienced and addressed will also have wider relevance for other environmental policies more broadly.

The aim of this study was to explore the barriers to and facilitators of the adoption, implementation and perceived effectiveness of takeaway management zone policies. We used a qualitative approach to explore local planning and public health government officers’ perspectives of these policies in England, UK.

## Methods

### Sampling and recruitment

In Autumn 2021, we identified 41 LAs that had adopted and implemented takeaway management zones. From those, we selected 28 LAs to make-up a diverse sample by region, deprivation (Index of Multiple Deprivation (IMD)) and variation in management zone policies. We also included LAs with unusual policy variations such as those with 200 metre or 10-minute walking distance management zones (as opposed to the more typical 400 metre approach as outlined earlier).

This final list of 28 LAs were approached and invited to participate in the study. As planning teams and public health teams are responsible for policy adoption and implementation within LAs, we initially identified senior managers from these teams and contacted senior managers via email to identify staff from their teams who could be interviewed about their work on takeaway management zones. Managers provided us with contact details of potential participants and we approached these individuals via email and invited them to participate. We also used snowballing techniques to recruit further participants. We continued recruiting participants until saturation was achieved, whereby no further new information related to the objectives of the present study were identified following preliminary analysis of the data (Fusch & Ness, 2015).

Of the 28 LAs approached, 15 participated and 13 either did not respond to the invitation (n=9) or declined due to lack of time (n=2) or lack of current involvement in the policy (n=2). LAs from the North (n=7), South (n=6), and West (n=2) of the country agreed to participate. From the 15 LAs who agreed to participate, we recruited 29 individual local government officers who worked in either planning roles (n=17), public health roles (n=10), or joint planning/public health roles (n=2). Participant characteristics are shown in Table 1.

**Table 1.** Participant characteristics.

| **Current Team/ Professional role** | **(n=29)** |
| --- | --- |
| <u>Public Health</u> |  |
| *Director/Head/Lead/Manager of Health & Wellbeing/Public Health* | 5 |
| *Public Health/Health Improvement Practitioner/Officer* | 3 |
| *Senior/Public Health Specialist* | 2 |
| <u>Planning</u> |  |
| *Lead/Principle/Head of Planning* | 1 |
| *Graduate/Senior/Planning Officer* | 8 |
| *Lead/Principle/Head/Manager of Policy Planning* | 3 |
| *Senior/Policy/Communications Officer* | 5 |
| <u>Combined Planning and Public Health</u> |  |
| *Head/Lead/Senior joint public health and planning role* | 2 |

### Data collection

Data collection took place between October 2021 and January 2022. Semi-structured one-to-one or paired (if requested by participants: n=3 pairs) interviews were conducted using videoconferencing software. Interviews were conducted in private spaces to maintain confidentiality and anonymity. Informed consent was sought before the interviews and participants had the opportunity to ask questions before, during and after the interviews.

Interviews were conducted using a topic guide for consistency, which was developed with the guidance of our study policy advisory group who had prior experience working in LAs and implementing planning policies. Interview questions broadly covered: professional background; policy background, adoption, and development; experiences of implementation; impacts of the policies; reactions from businesses and members of the public; the future of the policy. We piloted the topic guide to check the relevance of topics and understanding of questions. All interviews were audio-recorded and transcribed verbatim. The transcripts were checked against audio-recordings for accuracy and anonymised by removing identifiable content.

### Data analysis

Anonymised transcripts were stored and analysed using NVivo (Version 12 Plus) software. An inductive approach was taken throughout the analysis, whereby codes and themes were data-driven. Data familiarisation was achieved by reading transcripts and listening to audio-recordings. Data were then coded descriptively to represent emerging topics. The codes were developed through discussion with the research team. This process was iterative, with researchers continuously revising and adapting codes until they felt codes accurately represented the data. We triangulated responses from participants in different LAs, which allowed us to compare multiple perspectives. Once coded, the data were then analysed thematically (Clarke & Braun, 2017). Themes were identified by searching for commonalities, discordant views, and underlying meanings behind the derived codes. We actively presented common, atypical and contradictory opinions to ensure that different perspectives were represented. Themes were derived iteratively through discussion within the research team.

Ethical approval for the study was granted by the London School of Hygiene and Tropical Medicine Ethics Committee (Reference: 26337) in October 2021.

## Results

We identified four main themes concerning the adoption and implementation of takeaway management zones: professional identities; variations in adoption and implementation; establishing systems and ensuring clarity; perceived effectiveness. Collectively they cover the decision to adopt the policy and deciding what it will look like, matters of implementation including establishing systems, and interpretation of the policy in day-to-day practice. We found the adoption phase was important in shaping what was implemented and how.

### Professional identities

Effective working relationships between LA planning officers and public health officers were important for successful policy adoption and implementation. This was mediated by differing professional identities and priorities associated with their respective roles. These differences sometimes caused conflict and proved problematic. There were also occasions when professional values across planning and public health were aligned, creating cohesion and a greater acceptance of collaborative working.

When making the case for adoption, public health officers were primarily concerned with the negative health impacts of takeaways. Planners, by contrast, had a broader set of considerations to adhere to when weighing-up the possible costs and benefits of the policy. This included the economic benefits that new takeaways could provide. Whilst public health officers acknowledged that this variance in aspects of interest may have limited collaborative working, they sympathised with planners as having to balance competing agendas. Particularly in areas of economic deprivation, both public health and planning officers had complex roles. The professional responsibilities of planners were partly driven by fulfilling their LA’s strategic (non-health) priorities and sometimes the policy was perceived as conflicting with these priorities.

> *“It’s economic activity isn’t it and it’s commerce…we are low, on many sort of those economic matrix metrics… planning has got a wider purview in terms of what they’re looking at. It’s not just the public health agenda, which, obviously public health is pretty much exclusively looking at.”*

> (Policy & Public Health professionals, North of England)

In 2012, after 38 years in the National Health Service (NHS) (i.e., the UK’s publicly funded healthcare system), responsibility for public health was moved back into LAs. The history of interaction between public health and LAs, especially the legacy of the return of public health to LA control, has influenced collaborative working. Some participants suggested that this history caused resistance and distrust among planners around the public health agenda and collaboration with public health colleagues.

> *“…way back in 2014, I think 2013, 2014, when public health first came to the local authority, because obviously we used to sit with the Primary Care Trust. So we did have a little, let’s just say resistance….”*

> (Public Health officer, North of England)

To overcome this perceived disruption and resistance, and to encourage collaborative working, several LAs had recently started employing dual-trained officers, with specialisms in both planning *and* public health. Dual-trained officers have an understanding of the professional values and agendas of both professions. Participants explained that this helped to facilitate a partnership whereby goals were aligned and communication was improved.

> *“…my post sits between the [public health team], but I had probably 50% of my work plan is directed by the [planning team]. So a lot of my projects are planning and public health projects… dually trained across both teams …there’s certainly a lot of partnership working… a big part is trying to bring agendas together and whether that be in individual project teams or strategically through making sure sort of the agenda of both becomes common agenda…”*

> (Public Health & Planning professional, South of England)

In other cases, champions for this policy were within planning teams . These were often individuals with an understanding of the role of planning in public health and an acceptance of the benefits of the policy. Their role was to positively influence the perceptions of other planners and to work collaboratively with public health officers.

> *“…we’ve got, our planning colleagues, [name of colleague] in particular, has always played a very key role in what we call our healthy work partnership… she’s represented on that, she sees it as important, she understands the role that planning has… which I think helps as well.”*

> (Public Health *officer*, North of England)

Where planners and public health officers worked collaboratively, they reported that time spent learning about each other’s roles and the opportunity for public health professionals to get familiar with the planning system and planning terminology valuable. Planning officers also spent time preparing public health colleagues for planning processes including formal hearing sessions during appeals of rejected takeaway planning applications. This equipped public health teams with the necessary knowledge to attend hearing sessions and provide representations on behalf of the LA.

> *“I think we both learned a lot from each other, because I’m not a planner… so it has been interesting sort of learning and have an appreciation of the planning system as well… I don’t come from a planning perspective. So I had to have that appreciation… to look at… what parameters are there in terms of planning policy”*

> (Public Health officer, North of England)

In terms of influencing the decision to adopt the policy, the role of policy champions went beyond cross-departmental working. These individuals, alongside influential people like councillors and those in senior positions within planning and public health, could drive adoption and implementation. In fact, the importance of specific and engaged individuals in overseeing and advocating for the policy from adoption through to implementation was described as a key factor in the success.

> *“… we had a public health counsellor… they fully understood, supported…ultimately it’s for them to champion politically, and to get the rest of the members involved in supporting it..”*

> (Public Health & Planning, South of England)

> *“…the objectors really tried to challenge the robustness of our evidence base and it helped that our Director of Public Health (DPH) was actually able to defend it…”*

> (Policy Officer, South of England)

#### Variations in adoption and implementation

Not all LAs adopted full management zones (i.e., where takeaways are denied if falling within a certain distance from a school). Some varied their design to suit local needs and pressures. This resulted in variation in the specifications of the policy, such as adopting policies that involved restrictions on opening times rather than an outright rejection of new takeaway applications.

> *“…we would put a period of time when the hot food takeaway had to be closed and that was different for primary and secondary, acknowledging that secondary school children can come out at lunchtime but primary school children can’t… and that was the safest approach to take and one where we felt that we could defend our position… I think the problem we were facing was the link between one additional hot food takeaway and does it have any impact on a child’s obesity and that hadn’t… We didn’t want to get to the point where we [were] approved and [then] the first appeal we lost…”*

> (Policy officer, North of England)

In some cases, LAs adopted town centre exempt zones, where applications for new takeaways in overlapping town centres would not be subject to regulation because these are sites that generate income and create employment opportunities. This was particularly apparent in seaside towns and deprived areas where takeaways were considered important for maintaining the vitality of the high street and local economy. Paradoxically, these also tended to be areas with greater wider inequalities and higher concentrations of takeaways. Refusing takeaways could result in vacant retail units, which was considered highly damaging and undesirable by local government officers.

> *“…we felt that within those [Town] centres, it wouldn’t be acceptable from a planning point of view to say, you can’t operate from that centre, particularly in centres where we might have a lot of vacant units, we felt it’s better from a planning perspective to have it occupied, and it’d be a viable business rather than it just standing… empty… we would rather that the unit was occupied…provides local employment and things like that.”*

> (Planning officer, North of England)

> *“…we’ve also got a slightly more relaxed policy within our main town centres… given that [name of LA] is a traditional seaside resort as well. So fish and chips etc, are part of that, town centre…”*

> (Public Health & Planning professional, South of England)

As described in the previous section, planning and public health agendas did not always align. Planners were required to consider the possible economic impacts of any policies, health-based or otherwise. Management zones were felt to introduce the prospect of damaging the local economy and resulted in some LAs adding protective clauses, preventing implementation of the policy in some instances. For example, if a proposed takeaway was perceived as providing enough economic benefit, then the application would be permitted, and the clause would be enforced. This was particularly the case where retail units were vacant for long periods before being proposed as potential takeaways. In which case, filling a vacant unit would be considered beneficial to town vitality and this outweighed concerns over health.

> *“…they didn’t want was this policy to mean that an active use of a unit would be prevented because the only option of hot food takeaway… we had that caveat in that if you had been vacant for six months… you tried to market it to other uses, nobody wanted it other than hot food takeaway, then we would allow hot food takeaway….”*

> (Policy officer, North of England)

It is important to note that economic and town vitality concerns did not trump considerations of health in all LA contexts, especially those with larger public health teams and those that had deliberately cultivated a focus on prevention. In these settings, health, wellbeing and providing opportunities for members of the public to ‘thrive’ were all considered as important as economic priorities. Officers working in these LAs described an organisational culture that purposely positioned takeaways as harmful. Specifically, that they did not benefit their local economy, and they negatively impacted the vitality of areas and threatened opportunities for members of the public to ‘thrive’. These LAs tended to adopt and implement full management zones, without building-in variations to protect the local economy.

> *“… these [takeaways] didn’t represent particularly great uses… the organisation understands itself very well in terms of its overall impact on health and wellbeing…we now have…a commitment broadly as an organisation, where [name of LA] should be somewhere where people can thrive… we see ourselves, in part at least… doing what we can to help bring about those situations where people can do that.”*

> (Public Health officer, North of England)

#### Establishing systems and ensuring clarity

Within LAs, working to establish a formal process for applying takeaway management zone policies was seen as beneficial because it helped ensure consistency and acted as a means of reinforcing implementation. This process typically involved a designated individual taking on responsibility for identifying applications for new takeaways, ascertaining those responsible for providing responses to such applications (including public health, policy and planning officers), implementing the criteria within the policy, and reaching a final decision.

> *“…I’ll just go through the process of establishing for that location, like ticking against all the criteria, does it pass or fail? And then… go through the decision-making process…. what is the latest childhood obesity data, and I’ll have a physical link in that so you can go and check it and it makes it easy. So if somebody else was to pick up my job they could use that data easily.”*

> (Planning officer, North of England)

The importance of establishing formal processes and having systems in place, rather than relying on ad-hoc approaches to implementation, is particularly evident where LAs had no such processes. Lack of process has led to some takeaway applications being missed and the policy not being fully implemented, potentially leading to the policy not achieving its full potential in some settings. Having enough officers with designated roles to cover all aspects of implementation and making sure that all relevant officers were aware of the policy were vital for ensuring that the policy worked to its full potential.

> *“One of the kind of learnings… is to make sure that you have someone checking the lists… That role of checking lists…is really important, because the new policy officer, a new planning officer might not be aware it’s there. So they might miss that bit in the checklist, also it’s very busy people trying to balance a thousand and one other things.… I think we’ve had an elected member, a local councillor message saying, ‘we’ve seen this, this application’s come in, can you do something about it? I don’t want another hot food takeaway in my ward’… it had slipped through the net… our office is like… ‘I don’t know how I missed that one’.”*

> (Public Health professional, South of England)

If planning permission for a new takeaway was refused by a LA, applicants could appeal the decision. Participants explained that being able to overcome appeals and uphold rejections was largely contingent upon having established a robust and clear process for actively reviewing the quantity and quality of applications received and monitoring the total number of takeaway planning applications received and percentage rejected, at both first decision and after any appeal. This information helped officers assess whether the policy facilitated takeaway refusal and if not, assess alternative approaches.

> *“… we have records of every planning application we’ve had in, and then we would simply just note down, has it been approved? Has it been refused? Why was it refused? And then if there’s been an appeal, whether we won, or whether we lost the appeal, why? Like I said, it’s been quite easy, because we have actually refused all of the hot food takeaway policies, and we won all of our appeals.”*

> (Planning officer, North of England)

In addition to being dependent on having a formal process for reviewing new takeaway applications, the clarity of the policy also affected how it was interpreted and therefore implemented. Clarity in the wording and the use of objective tools such as maps and buffer areas were important in ensuring that planners could apply the policy with confidence. This reduced the likelihood of refusing applications that would be subsequently overturned on appeal and, in the process, generate expense and extra workload because the policy had been inaccurately measured.

> *“… Are the criteria clear to them as in what they can allow and refuse? Or is it a policy that they see is woolly… with the appropriate mapping, for the existing uses and the buffer areas, it’s relatively easy for them to implement and assess.… it’s because if they can be more confident about refusing an application… and so they think they can basically sort of be more objective with why they’re refusing it.”*

> (Policy officer, South of England)

#### Perceived effectiveness

Participants explained that in most cases, they thought the policy had been effective in denying permission for new takeaways. Some participants also reported that the LAs within which they worked no longer received takeaway applications near schools and, therefore, they no longer had to use the policy as much as they previously did. They implied that this may be because having the policy in place deterred the submission of new takeaway applications, or perhaps that prospective takeaway operators had simply found alternatives ways of operating.

> *“… the benefits… being we have had success in it, we have had refusal. So the benefit has been less hot food takeaways than would have been without it …”*

> (Public Health officer, South of England)

> *“…it’s dried up at the moment, I haven’t had a hot food enquiry for about six months now, a year. We went through a spate where there was quite a few but whether people now are looking to get round it the other way by opening up a set of industrial units and having [food] delivered which beats… this particular system.”*

> (Planning officer, West of England)

In the quote above, the participant refers to setting up *“industrial units and having [food] delivered”*. These types of operations are commercial kitchens with delivery centres, sometimes known as “dark kitchens”. Rather than being customer-facing retail takeaways, the sole purpose of these establishments is to prepare meals for delivery. Typically, converting retail or restaurant space to a commercial kitchen does not require planning permission, if there is no external building work needed (Howard Kennedy LLP, 2022). In which case, dark kitchens can be a more efficient than customer-facing retail and a less regulated commercial operation.

In addition to dark kitchens, participants explained that businesses were finding other new ways to operate. For example, restaurants and cafés could also operate as takeaways if this was classed as a secondary, or ‘ancillary’, function. The threshold for this varies between LAs and tends to be calculated on percentage of floorspace devoted to the secondary function(s) and the percentage of profits it generates. Without exceeding this threshold, an outlet could still sell takeaway-type food and offer a takeaway services but might be primarily classified as a restaurant.

> *“I guess the main limitation is applications coming in for cafés and restaurants with a takeaway use, they say is ancillary [secondary function]. …the way around it would be to say that you’re a restaurant or a café and include a seating area and say that the takeaway is ancillary… [name of takeaway outlet]… they’ve successfully proven that they are a restaurant café, and that’s now accepted.”*

> (Policy officer, South of England)

Almost all participants reported that although they believed the policy facilitated the rejection of new takeaways, it did not tackle the unhealthy local food environment more generally. For example, the impact of existing takeaways, and new and existing sweet shops, bakeries and restaurants, which also sell unhealthy foods. It was argued that this was due to the planning infrastructure and the narrow definition of a takeaway, which limited the role of planning in effectively regulating the food environment.

> *“…it only scratches the surface… It has no restriction on all other unhealthy food outlets…having kids going to the corner shop [convenience store] buying handfuls of unhealthy food… can’t possibly restrict… shop uses, because we need shops…”*

> (Planning officer, North of England)

> *“… this is only about a request for a licence for an application for a new A5. So all the existing ones… we need to do something about the volume.”*

> (Public Health officer, South of England)

## Discussion

### Summary of main findings

In this study we explored takeaway management zones around schools in England, UK with respect to barriers to and facilitators of adoption, implementation, and perceived effectiveness. We used a qualitative approach to study local government officers’ perspectives of the policies. Our findings suggest that effective working relationships between LA planning and public health officers were important for policy adoption and implementation. Concerns over the possible economic impacts of the policy sometimes caused tensions between these two groups at adoption and implementation. LAs reporting successful adoption and implementation took steps to resolve these tensions by developing roles across both departments and appointing policy champions. Not all LAs adopted full management zones and some made adaptations and compromises based on what they felt was supported by evidence, as well as to protect town centre vitality and the local economy. Working to establish a formal process for implementing management zone policies and clarity in the policy wording was seen as beneficial because it helped ensure confidence and consistency and reinforced implementation. There was a perception that the policy had facilitated the rejection of applications for new takeaways and, in some cases that LAs had received fewer or no further new takeaway applications near schools. Although this was viewed as positive, it was noted that some businesses might have found new ways to operate that avoided such policies.

### Strengths and limitations

To the best of our knowledge, this is the first process evaluation of the adoption and implementation of takeaway management zones around schools in England and takeaway restriction policies internationally. We employed a rigorous approach to data analysis, whereby transcripts, codes and themes were continuously discussed by the analytical team in data clinics and amended iteratively until the written presentation of the findings to ensure they captured the data as accurately as possible. We collected and compared responses from officers working in LAs with different area and policy characteristics across England. As such, our findings are potentially transferable to other LAs adopting and implementing these policies in England or similar policies elsewhere regardless of their area characteristics.

However, the officers who participated in the study were drawn from a sample of LAs that may not necessarily be representative of all LAs adopting or implementing management zones and those choosing not to adopt policies. Approximately half the LAs we approached (13/28) refused participation and it may be that those who participated were more motivated, involved and engaged in adopting and implementing the policy. We also interviewed more officers from the planning profession (17/29) as opposed to those in public health (10/29). This may have resulted from the latter’s involvement in the Covid-19 pandemic response. However, it is unlikely that this impacted the data collected as public health officers experiences and perceptions were mostly consistent with those of their planning colleagues. Lastly, in focusing exclusively on the accounts of local government officers and their decision-making, we have not addressed the wider political contexts in which these policies operate and the role of elected members or the planning inspectorate.

### Contributions to knowledge and practice

Food businesses play a critical role in shaping the food environments of individuals and populations. The concept of the commercial determinants of health has been used elsewhere to focus attention on the role of food and beverage companies as important drivers of non-communicable diseases globally (Gomez et al., 2024; Maani et al., 2020). These commercial determinants of health include food business practices, which create conditions that increase the availability, accessibility, and consumption of highly unhealthy foods (Chung et al., 2022), In Europe, there is a gap in food retail environment policies despite the global focus on targeting food environments particularly surrounding children (Pineda et al., 2022; World Health Organisation (WHO), 2022). Takeaway management zones around schools restrict the proliferation of takeaways when implemented and, in doing so, constrain the growth in availability and accessibility of takeaway foods to children and the general population more broadly (Rahilly et al., 2024). Unhealthy food environments may be especially detrimental to children, which strengthens the case for their adoption (Soon et al., 2023). Such policies can serve to disrupt the balance of power that currently favours commercial interests over public health (Chung et al., 2022). This is supported in our findings where in some cases LAs were no longer receiving applications since the policy had been adopted and implemented; the policy may have acted as a deterrent through serving as an additional barrier that prospective takeaways would have to overcome. Our findings may therefore encourage other countries to adopt and implement similar policies to improve local food environments and be used as a basis to overcome issues related to adoption and implementation.

Planning officers roles require the need to consider a range of factors, notably economic benefits. However, management zone policies tend to originate in public health departments which can result in the framing of these policies as a potential threat to planners. This has been reported in other work (Chang & Radley, 2020; Keeble et al., 2021; Lake et al., 2017; Pineo & Moore, 2021), including previous qualitative work on local government officers’ experiences of adopting planning regulations addressing takeaways (Keeble et al., 2021). Our findings revealed tensions and conflicts, broadly speaking between economic development and public health agendas. There was evidence that policy decisions were made in economic contexts, with protective clauses to avoid vacant retail units and permit takeaways if perceived to negatively affect economic development. Pursuing economic development goals means that LAs engage closely with food businesses in order to generate revenue and boost local investment (McKevitt et al., 2023). Commercial actors can seek to influence LAs and resist regulations by terminating local investments and resources if new public health policies are introduced and by litigating against public health polices, like takeaway management zones (McKevitt et al., 2023). Despite these challenges, there is a need to develop effective ways to better balance economic and health priorities. Measures designed to help populations lead healthier lives need to be embedded within economic and development strategies, and local businesses must be held accountable for their impact on community health (Chung et al., 2022). Participants in the present study recounted examples of LA approaches to cultivating a focus on health and positioning it as equally important as the local economy. Alternative retail spaces that are not takeaways could be more strongly considered to create economic development.

It was also speculated that businesses found alternative ways to circumvent the policy, such-as locating dark kitchens outside takeaway management zones or masquerading as restaurants with seating areas. This suggests that other policies with a broader focus on improving population health targeting different unhealthy food sources in the retail food environment are needed. Countries adopting and implementing such strategies should be supported by central government to achieve this (Gomez et al., 2024).

Previous research has also reported strained working relationships between planning and public health colleagues owing to differences in knowledge, priorities, perception of role, training, professional silos and lack of resources (Carmichael et al., 2013; Carmichael et al., 2012; Chang & Radley, 2020; Ige-Elegbede et al., 2021; Lake et al., 2017). We found that staff with training in or experience of both public health and planning, or local policy champions helped to bridge the gap between planning and public health, facilitating adoption and aiding implementation. The value of interdepartmental and interdisciplinary training to collaborative working has also been highlighted elsewhere (Lake et al., 2017). However, LAs often lack resources and staff to do this (Carmichael et al., 2013; Carmichael et al., 2012; Chang & Radley, 2020; Ige-Elegbede et al., 2021). Alternatively, external funding for qualifications (e.g. Masters, PhDs etc.) in public health or planning from national funding bodies such as the UK National Institute for Health Research (NIHR) may be sourced.

Our analysis builds understanding of the decision-making processes of planners when they review takeaway planning applications and what may help them implement related policies. In this study, planners valued the use of objective measures such as buffer maps to clearly define takeaway management zones, clarity in the wording of the policy and established internal processes for reviewing applications. This suggests a need to involve and/or gain feedback from officers responsible for implementing such policies internally to ensure consistency and confidence in implementation. In our study, a lack of research evidence demonstrating health benefits and evidence weighing up the economic costs and benefits led to diluted policies. The gap between researchers and local decision makers has also been identified as a barrier to using research evidence in public health decision making (Orton et al., 2011).

### Conclusions

Interventions to address diet related non-communicable diseases remain a public health priority globally. In England, LAs are tasked with designing, adopting, and implementing policies to address this disease burden, whilst also managing their social, economic, and environmental impacts. Takeaway management zones around schools are intended to modify the how the local food environment will develop in the future by restricting the future proliferation of takeaways in these areas, and thereby curbing children’s exposure to them. Once adopted, the implementation of these policies is affected by factors both internal and external to the LAs delivering them. Food environments are also retail environments, which can bring planning professionals into conflict with their public health colleagues. However, despite their sometimes divergent agendas, our findings suggest that when planners and public health officers worked collaboratively, their complementary skills proved beneficial in designing and implementing a policy they perceived to be effective. This perceived effectiveness of takeaway management zones around schools in most cases, suggests the policy can achieve system change. Whilst new businesses may find alternative ways to operate, takeaway management zones may act as a disincentive to takeaway planning applications and an incentive for businesses to sell healthier options. This policy demonstrates what can be achieved through partnership working. The lessons learned from our study can be built upon globally and adapted to the changing food purchasing landscape to achieve changes in the food environment.

## Data Availability

All data produced in the present work are contained in the manuscript.

## Acknowledgements

We would like to thank all the participants who kindly gave up their time to take part in the interviews.

## Funding

This study was funded by the National Institute for Health Research (NIHR) Public Health Research Programme (Project number: NIHR130597). The views expressed are those of the author(s) and not necessarily those of the NIHR or the Department of Health and Social Care. MK, MW, JA and TB were supported by the Medical Research Council (grant number MC_UU_00006/7). OM was supported by a UKRI Future Leaders Fellowship (MR/T041226/1). CT was supported by the NIHR Applied Research Collaboration (ARC) East of England. For the purpose of open access, the authors have applied a Creative Commons Attribution (CC BY) licence to any Author Accepted Manuscript version arising.

## Notes

### Competing Interest Statement

The authors have declared no competing interest.

### Author Declarations

Ethics committee of London School of Hygiene and Tropical Medicine gave ethical approval for this work.

